# Nationwide SARS-CoV-2 Surveillance Study for Sewage and Sludges of Wastewater Treatment Plants in Turkey

**DOI:** 10.1101/2020.11.29.20240549

**Authors:** Bilge Alpaslan Kocamemi, Halil Kurt, Ahmet Sait, Hamza Kadi, Fahriye Sarac, Ismail Aydın, Ahmet Mete Saatci, Bekir Pakdemirli

**Affiliations:** Marmara University, Department of Environmental Engineering, Istanbul, Turkey; Saglik Bilimleri University, Hamidiye International Scholl of Medicine, Department of Medical Biology, Istanbul, Turkey; Ministry of Agriculture and Forestry, Republic of Turkey, Veterinary Control Central Research Institute, Pendik, Istanbul, Turkey; Ministry of Agriculture and Forestry, Republic of Turkey, Veterinary Control Central Research Institute, Samsun, Turkey; Turkish Water Institute, Istanbul, Turkey; Ministry of Agriculture and Forestry, Republic of Turkey, Ankara, Turkey

**Keywords:** Covid-19, primary sludge, polyethylene glycol, RT-qPCR, SARS-CoV-2, virus concentration, wastewater, waste activated sludge

## Abstract

Since the announcement of the pandemic of Covid-19 by WHO on March 11, 2020, the countries have started to monitor surveillance of SARS-CoV-2 through medical tests. However, people with no and very light symptoms are usually not medically tested or never hospitalized and they are missed. In the study of Wu et al. [1], it was realized that the urine and faeces of all infected people contain SARS-CoV-2. After that, sewage, and sludge-based SARS-CoV-2 surveillance studies have gained significant importance around the world (Fig.1). SARS-CoV-2 was detected in wastewaters in The Netherlands [2,3,4], USA [1,5,6,7, 8, 9, 10], Australia [11], France [12, 13, 14], China [15], Spain [16,17,18,19,20], Italy [21, 22,23], Israel [24], Turkey[25], Germany[26], Japan [27,28], India [29,30], Pakistan [31], Brazil [32,33], Chile [34], Denmark, France, Belgium[35], Equator [36] and Sweden [37] using different virus concentration techniques. Published data show that high concentrations of the SARS-CoV-2 RNA reaches to wastewater treatment plants (WWTPs). On 7^th^ of May 2020, Turkey took its place among a few country which have been started wastewater based surveillance studies at the early stages of pandemic by reporting SARS-CoV-2 RT-qPCR levels of major WWTPs of Istanbul [25]. Turkey [38] first detected SARS-CoV-2 in both primary and waste activated sludges of Istanbul WWTPs. Later, USA [39] and Spain [40] were also studied on sludge samples. There are also studies evaluating the SARS-CoV-2 in WWTPs effluents [10,13,14, 28, 29,30, 34, 36].

This study aimed to scan distribution of Covid-19 through Turkey by SARS-CoV-2 measurements in influent, effluent and sludge samples of WWTPs. The influent, effluent and sludge samples were collected from main WWTPs located in 81 cities of Turkey through May 2020-July 2020. Among those 81 cities, Istanbul metropole with 15.5 million inhabitants was chosen as the pilot city since 65% of all cases in Turkey were present here. Hence, all treatment plants in Istanbul were scanned through the study. The viral activity tests were also conducted for the influent, effluent and sludge samples resulting high qPCR.

## 2. Value of the Data

- The dataset demonstrates a nationwide SARS-CoV-2 surveillance study which is unique worldwide. It is the first demonstration of evaluating Covid-19 distribution in a whole country of 83 million through a SARS-CoV-2 surveillance study performed in wastewater.
- The dataset provides information about fate of SARS-CoV-2 in wastewater treatment plants by demonstrating SARS-CoV-2 levels in influent, effluent, primary sludge and waste activated sludge samples.
- The dataset provides information about the possible activity of SARS-CoV-2 in various parts of wastewater treatment plants, as a first survey in the world

## 3. Methodology

### 3.1. Sampling

In Turkey, there are 3 major types of wastewater treatment plants. Type 1, called as preliminary treatment plants include physical treatment units, screens and grit chambers. Type 2, called as activated sludge plants include physical treatment units and biological organic carbon removal and nitrification units. Type 3, called as advanced wastewater treatment plants include physical treatment units and biological carbon, nitrogen and phosphorus removal facilities. There also a few cities not including any treatment facility for domestic wastewater. Some of the Type 2 and type 3 plants do not have primary sedimentation. Thus, since there is no primary sludge, there are no anaerobic digesters and the waste activated sludge (which is mostly bacteria) is stabilized by extended aeration.

In May 2020-June 2020, influent, effluent, primary and waste activated sludge samples were collected from 189 WWTPs of 81 cities of Turkey (Figure 2). Totally, 177 influent, 167 effluent, 35 primary sludge and 122 waste activated sludge samples were collected. In megacity Istanbul where about 65% cases were recorded, the samples were collected from 25 treatment plants (7 preliminary treatment plants, 9 activated sludge plants, 9 advanced activated sludge plants) in order to observe the areal distribution of the cases. For the other cities, treatment plants serving highest population were selected for sampling. In the big cities, more than one treatment plant was selected depending on the population of the city. In few cities not having any wastewater treatment, the samples were collected from the manholes located at center of the city.

**Fig 1.**
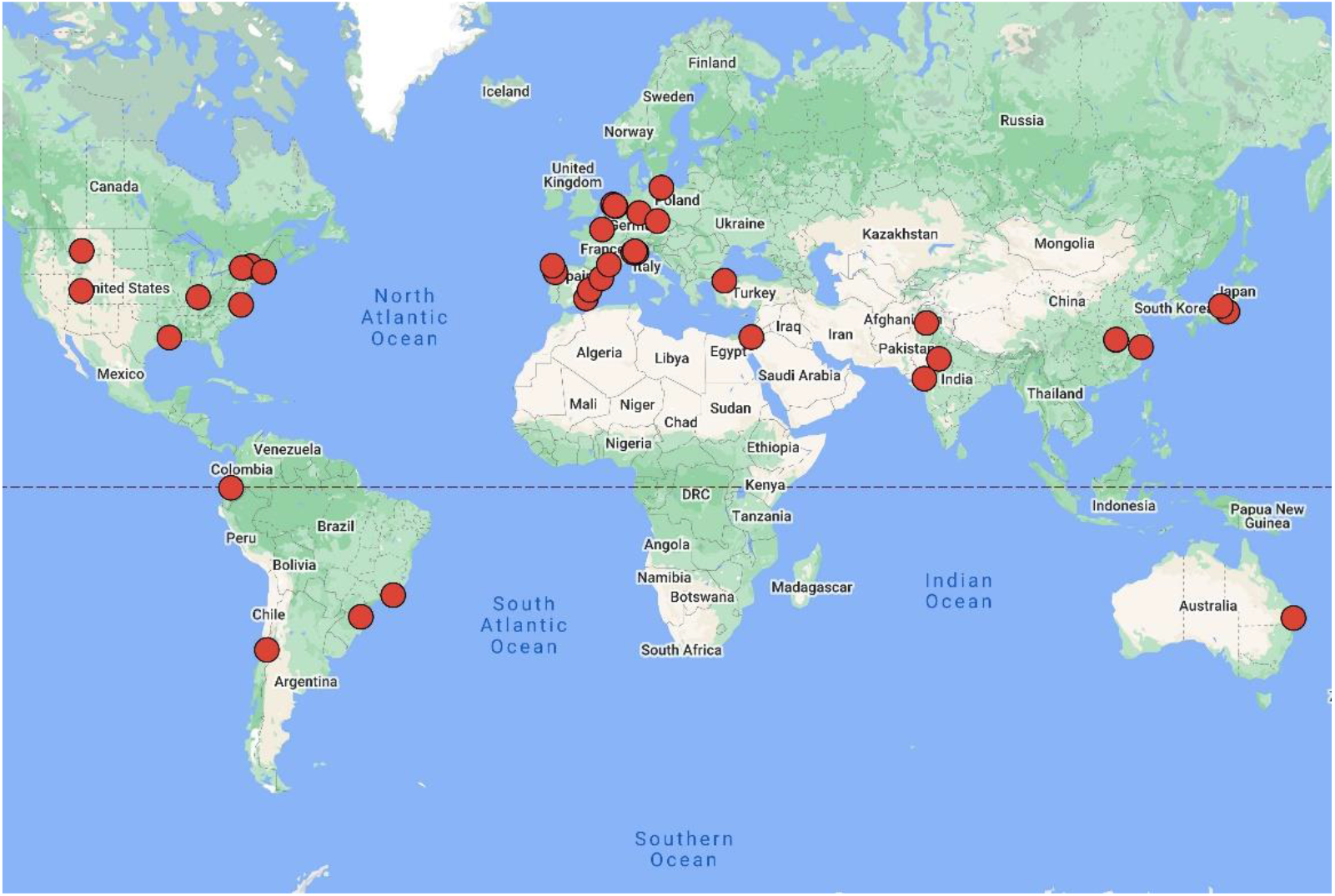
Worldwide SARS-CoV-2 surveillance studies in wastewater (https://www.google.com/maps/)

**Fig 2.**
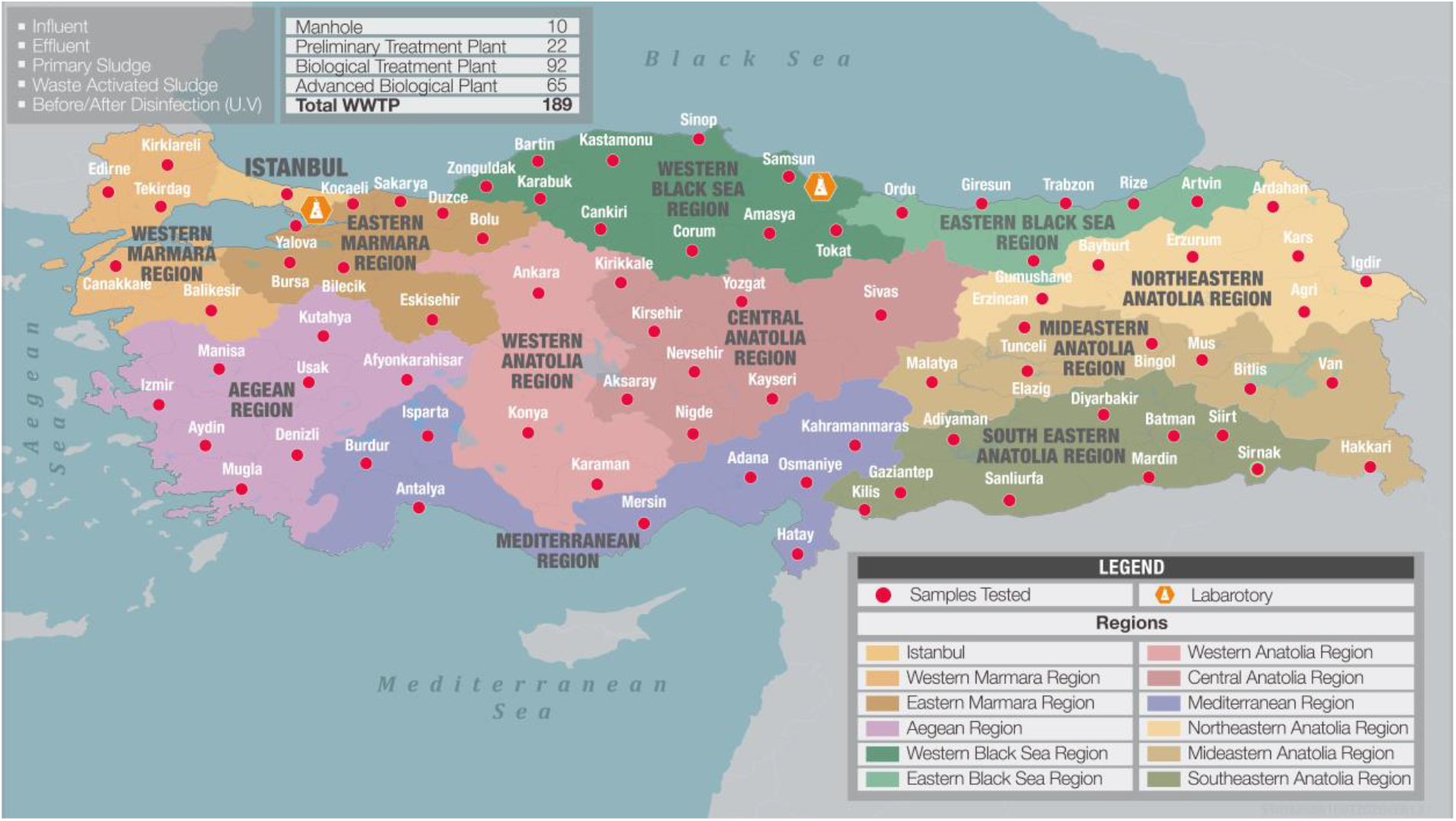
Sampling locations of nationwide SARS-CoV-2 surveillance study in wastewater and sludges of Turkey.

The samples collected in 250 ml sterile bottles were transferred under cold chain (less than 4 degrees Celsius) either to Istanbul Pendik or Samsun Veterinary Control Central Research Institute depending on the location of the city.

### 3.2. SARS-CoV-2 concentration

Samples were shaken at 4°C at 100 rpm for 30 min in order to transfer attached viruses to the aqueous phase. Microorganisms and large particles were removed from the samples by centrifugation at 7471G for 30 minutes at 4°C. 250 ml of supernatant was filtered through 0.45 μm and 0.2 μm to remove remaining particles and cell debris. Filtrate was mixed thoroughly with PEG 8000 (10% w/v) by shaking for 1 minute. The mixture was incubated at 4°C at 100 rpm for overnight. Following to the incubation, the mixture was divided in six 50 ml falcon tubes. Viruses were precipitated by centrifugation at 7471G for 120 minutes at 4°C. Supernatant was removed carefully without disturbing the pellets. Pellets of each falcon tubes were re-suspended with 200 μl RNA free water. 1 ml of virus concentrate was used for total RNA extraction and remaining concentrate stored at -80°C. Total viral RNA was extracted either with Roche MagNA pure LC total nucleic acid isolation kit using Roche MagNA pure LC system in accordance with the manufacturer’s protocols (Istanbul Pendik Lab) or manual RNA extraction method (Samsun Lab). RNA was determined both qualitatively and quantitatively by Thermo NanoDrop 2000c (Penzberg, Germany). Due to high community risk of using SARS-CoV-2 virus, 300 µl of 10^5^ copy/ml surrogate avian coronavirus of Infectious Bronchitis Virus were added our samples in order to evaluate the virus recovery efficiency of PEG 8000 adsorption method. Based on RT-QPCR results, 1-1.5 log virus titer loss were observed after PEG 8000 adsorption and RNA isolation.

### 3.3. Quantitative reverse transcription PCR (RT-qPCR)

Primers and taqman probe sets targeting SARS-CoV-2 RdRp gene were used in this study (Corman et al., 2020) to detect and quantify SARS-CoV-2 virus. Serial dilution of synthetic SARS-CoV-2 RdRp gene were used as a standard for absolute quantification. RT-qPCR analysis was performed in Realtime ready RNA virus Master (Roche Diaonostics, Mannhaim, Germany) contained 0.8 nM of forward primer and reverse primer, 0.25 nM probe and 5 μL of template RNA. The RT-qPCR assays were performed at 50 °C for 6 min, 53 °C for 4 min, 58 °C for 4 min for reverse transcription, followed by 95 °C for 1 min and then 45 cycles of 95 °C for 10 s, 58 °C for 30 s and 72 °C for 1 s for data collection using a Roche LightCycle 2.0 thermal cycler (Roche Diaonostics, Mannhaim, Germany).

### 3.4. SARS-CoV-2 Activity Tests

Samples which viral nucleic acid detected by RT-QPCR were filtered through 200 um filter prior to inoculation. 200 µl of samples were inoculated to Pendik Veterinery Research Control Institute vero cell line into 24-well plates, shaken every 15 minutes and kept at 37 °C for 1 hour, waiting for adsorption. After adsorption, the liquid in the wells was drawn and added to 1000 µl Glasgow Minimum Essential Medium (GMEM) which containing 2% fetal calf serum, 1% antibiotic and L-Glutamine. It was examined in terms of cytopathic effect (CPE) with a microscope for 3 days. The second passages were inoculated on 24-well tissue culture plates in the same order by the adsorption method. 200 µl from the first passage was inoculated into 24 well plates. It was surveyed in terms of CPE again. The third passage was inoculated using same method.

## 4. Data Description

### 4.1. SARS-COV-2 RT-qPCR Results

The SARS-CoV-2 levels (virus titer/L) detected in WWTPs of 81 cities of Turkey are demonstrated due to geographical regions of Marmara (*Istanbul, Fig.3; Eastern Marmara, Fig.4*; *Western Marmara, Fig.5*), Anatolia (*Western Anatolia and Central Anatolia, Fig.6*), Aegean (*Fig. 7*), Black Sea (*Western and Eastern Black Sea, Fig.8*), Mediterranean (*Fig. 9*), Eastern Anatolia (*Northeastern and Mideastern Anatolia, Fig. 10; Southeastern Anatolia, Fig.11)* regions.

**Fig 3.**
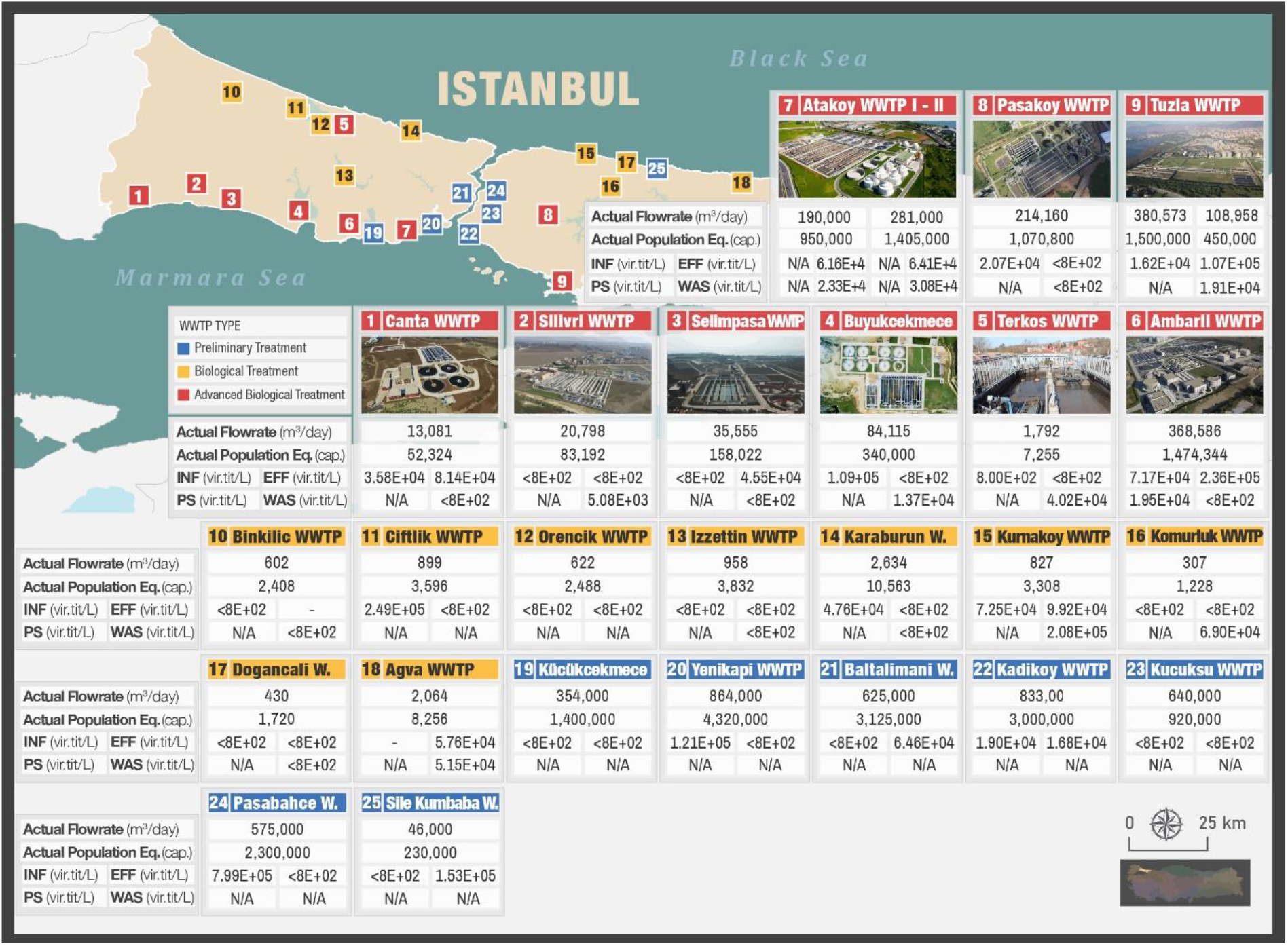
SARS-CoV-2 titers in wastewater and sludges of Istanbul WWTPs

**Fig 4.**
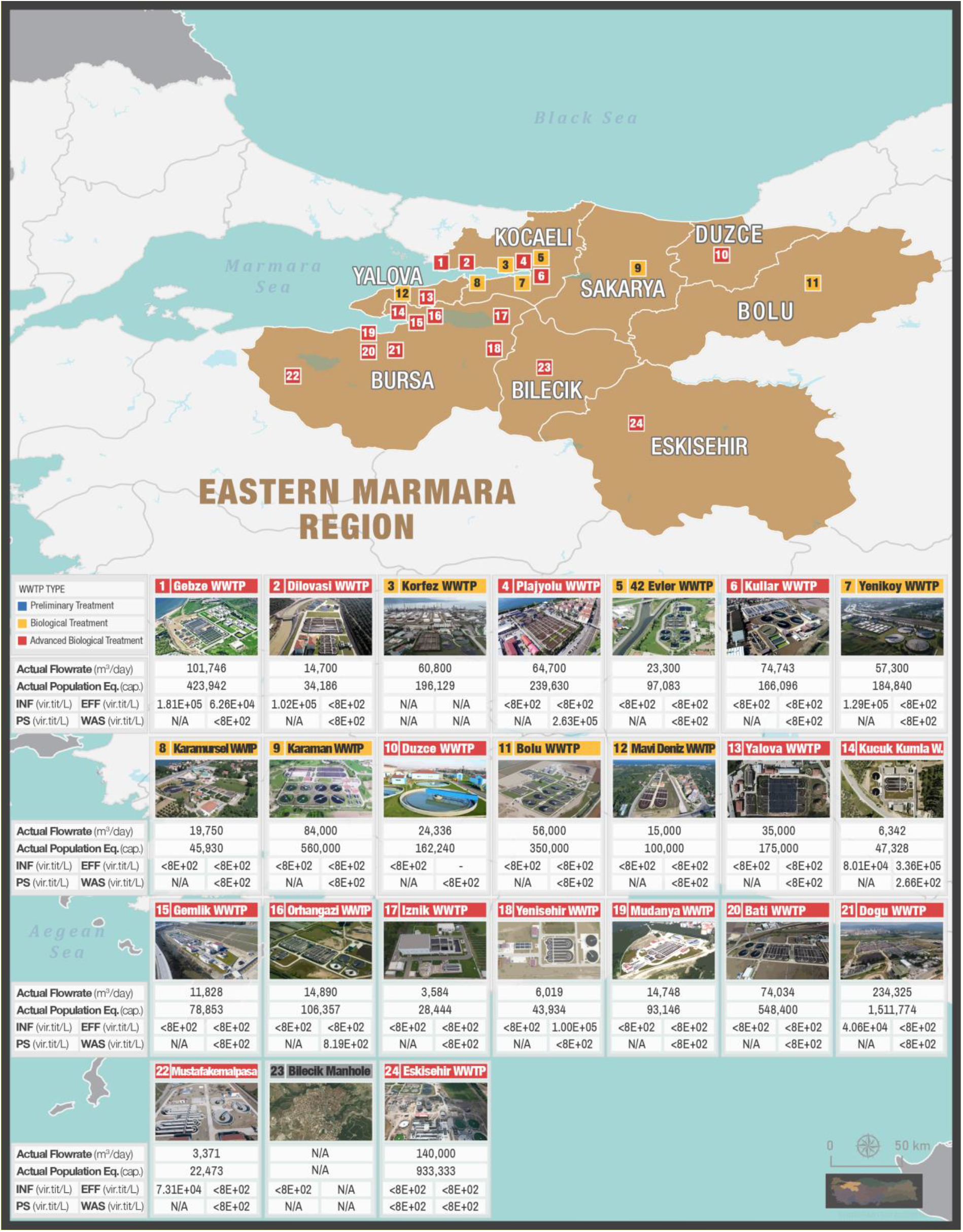
SARS-CoV-2 titers in wastewater and sludges of cities located in Eastern Marmara region

**Fig 5.**
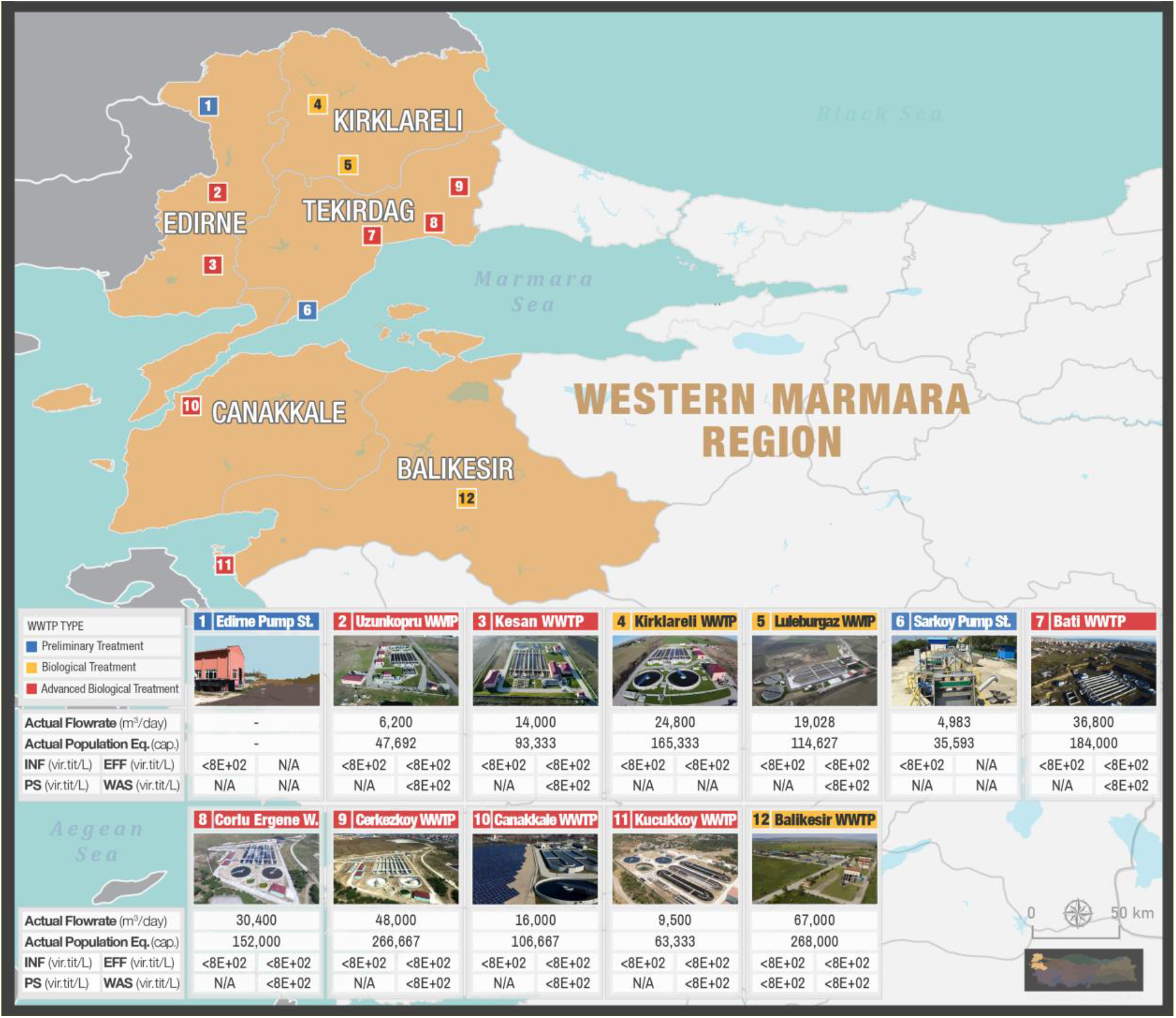
SARS-CoV-2 titers in wastewater and sludges of cities located in Western Marmara region

**Fig 6.**
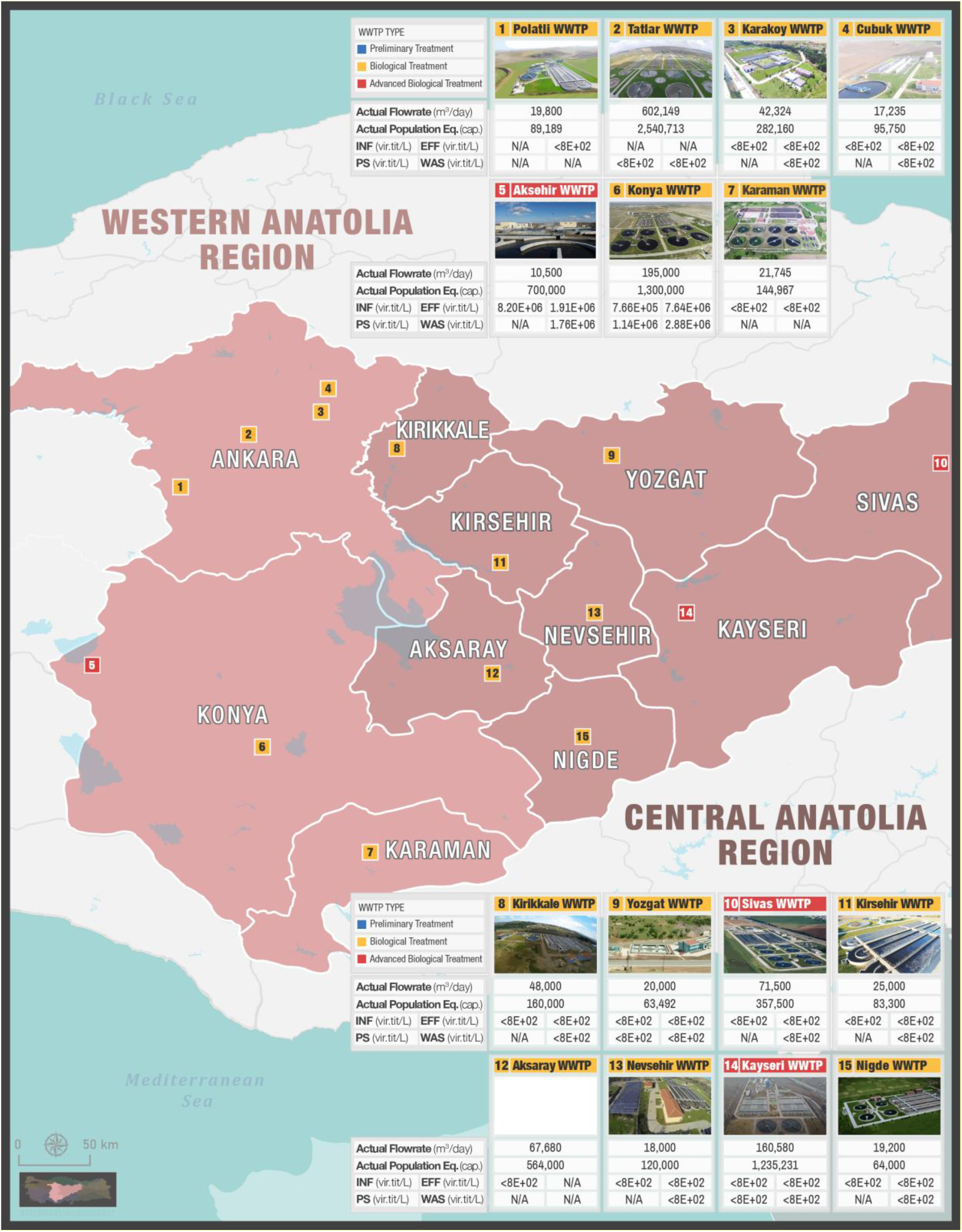
SARS-CoV-2 in wastewater and sludges of cities located in Western and Central Anatolia region.

**Fig 7.**
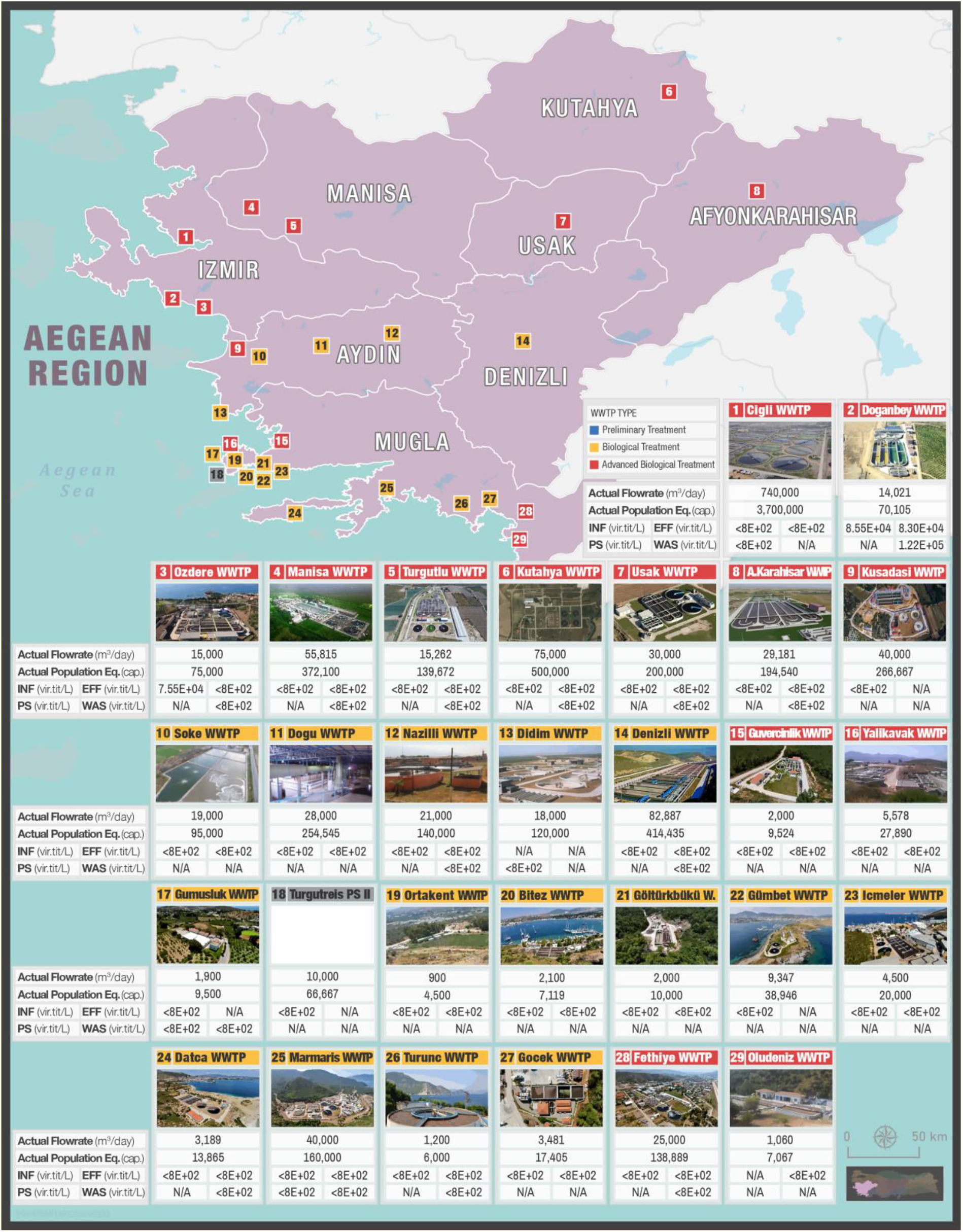
SARS-CoV-2 in wastewater and sludges of cities located in Aegean region.

**Fig 8.**
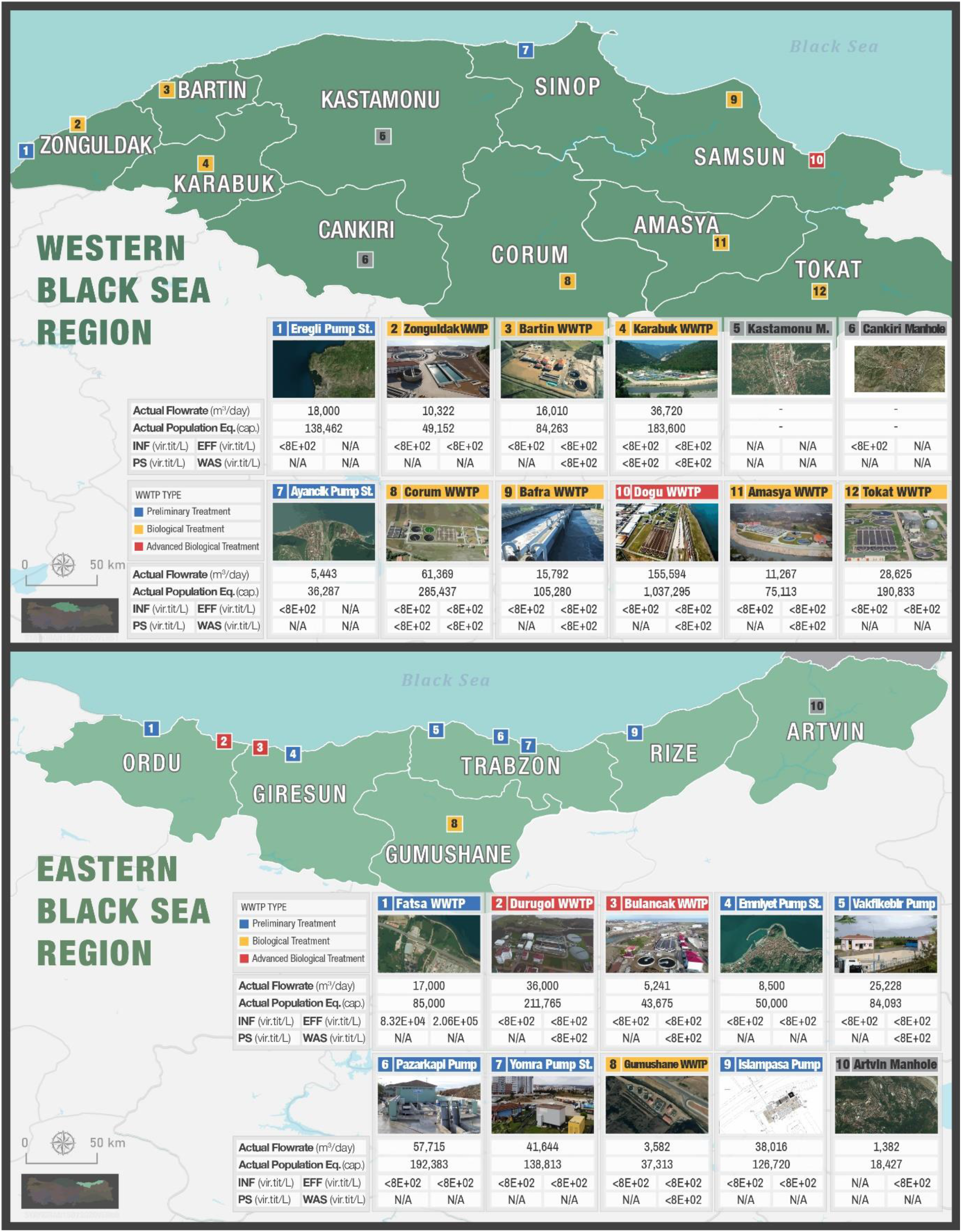
SARS-CoV-2 titers in wastewater and sludges of cities located in Black Sea region

**Fig 9.**
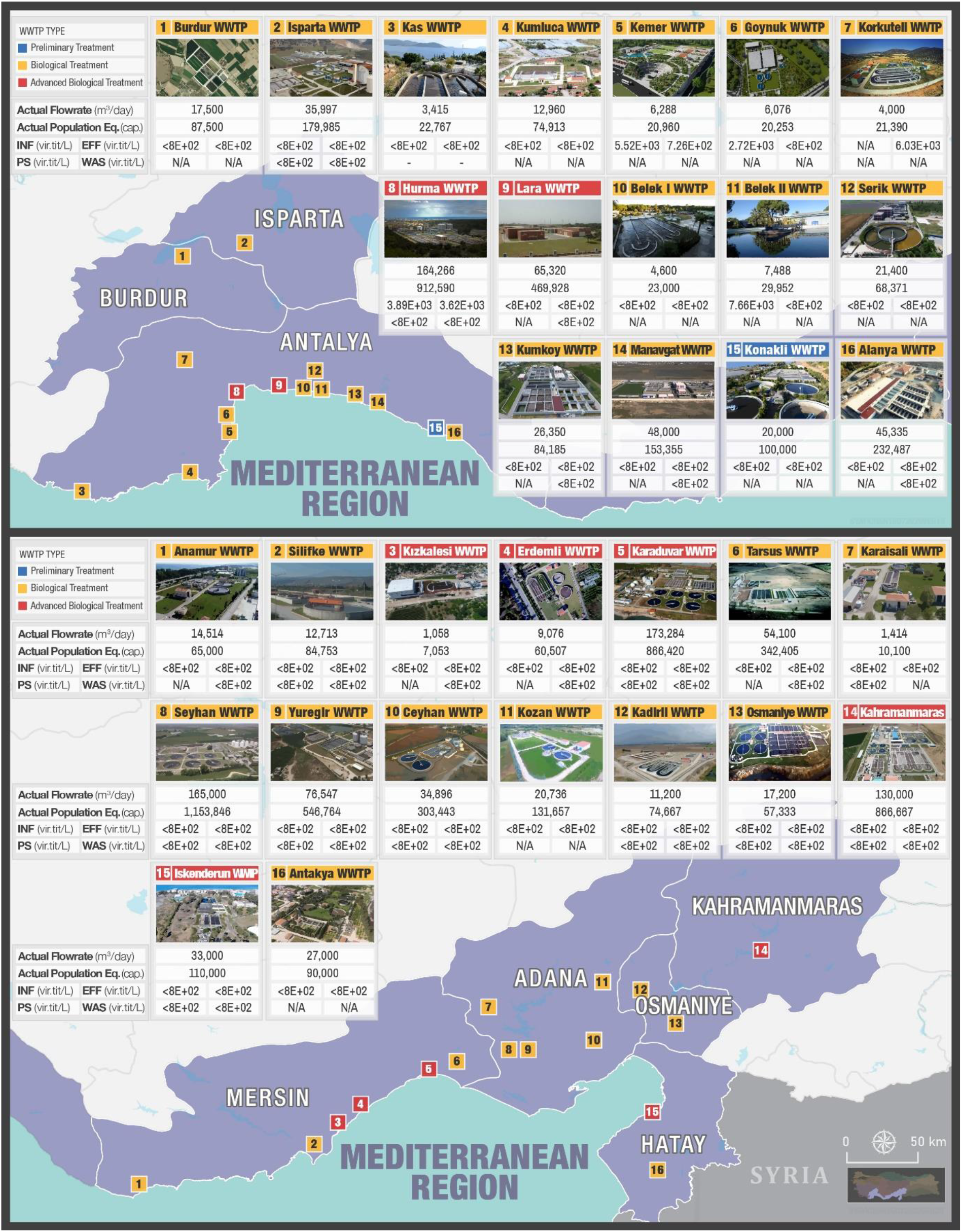
SARS-CoV-2 titers in wastewater and sludges of cities located in Mediterranean region

**Fig 10.**
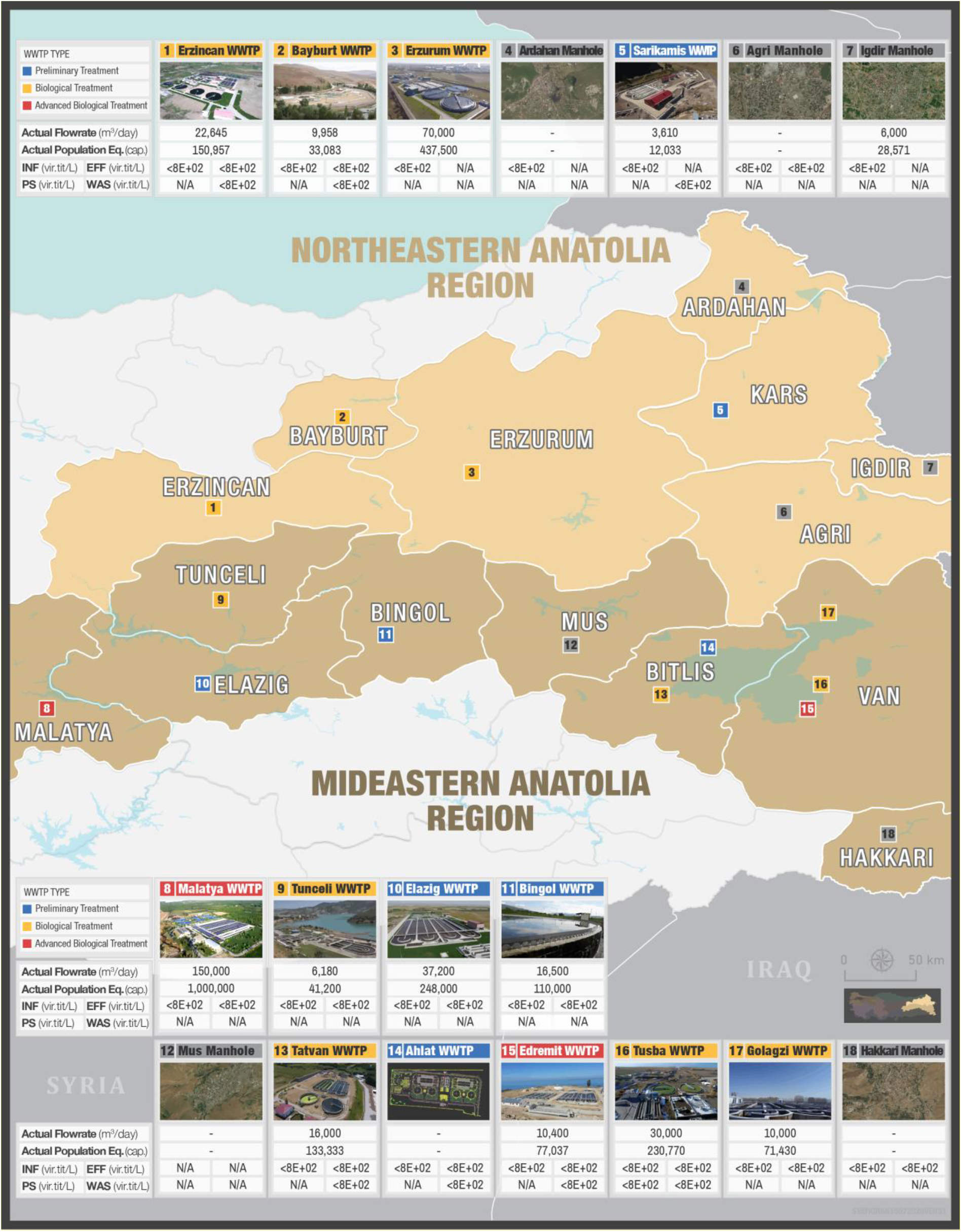
SARS-CoV-2 titers in wastewater and sludges of cities located in Eastern Anatolia region

**Fig 11.**
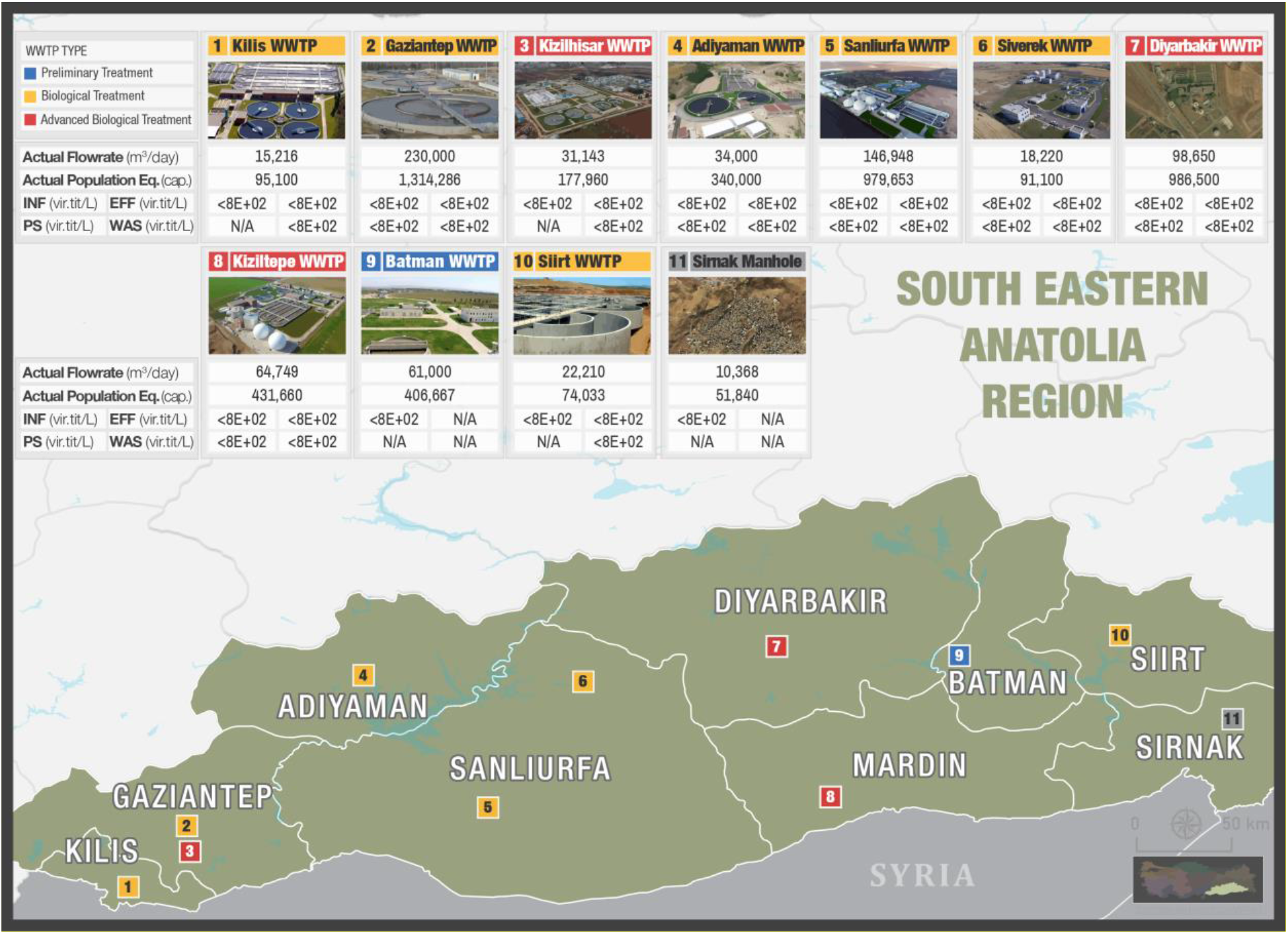
SARS-CoV-2 titers in wastewater and sludges of cities located in SouthEastern Anatolia region

SARS-CoV-2 genome was detected quantitatively in Western Marmara (Istanbul), Eastern Marmara (cities of Bursa and Izmit), Aegean (city of Izmir), Eastern Black Sea (city of Ordu), Mediterranean (city of Antalya) and Central Anatolia (city of Konya) regions in the range of 8 E+3-8E+6 virus titer/L. These results are summarized on Turkey map shown in Figure 12.

**Fig 12.**
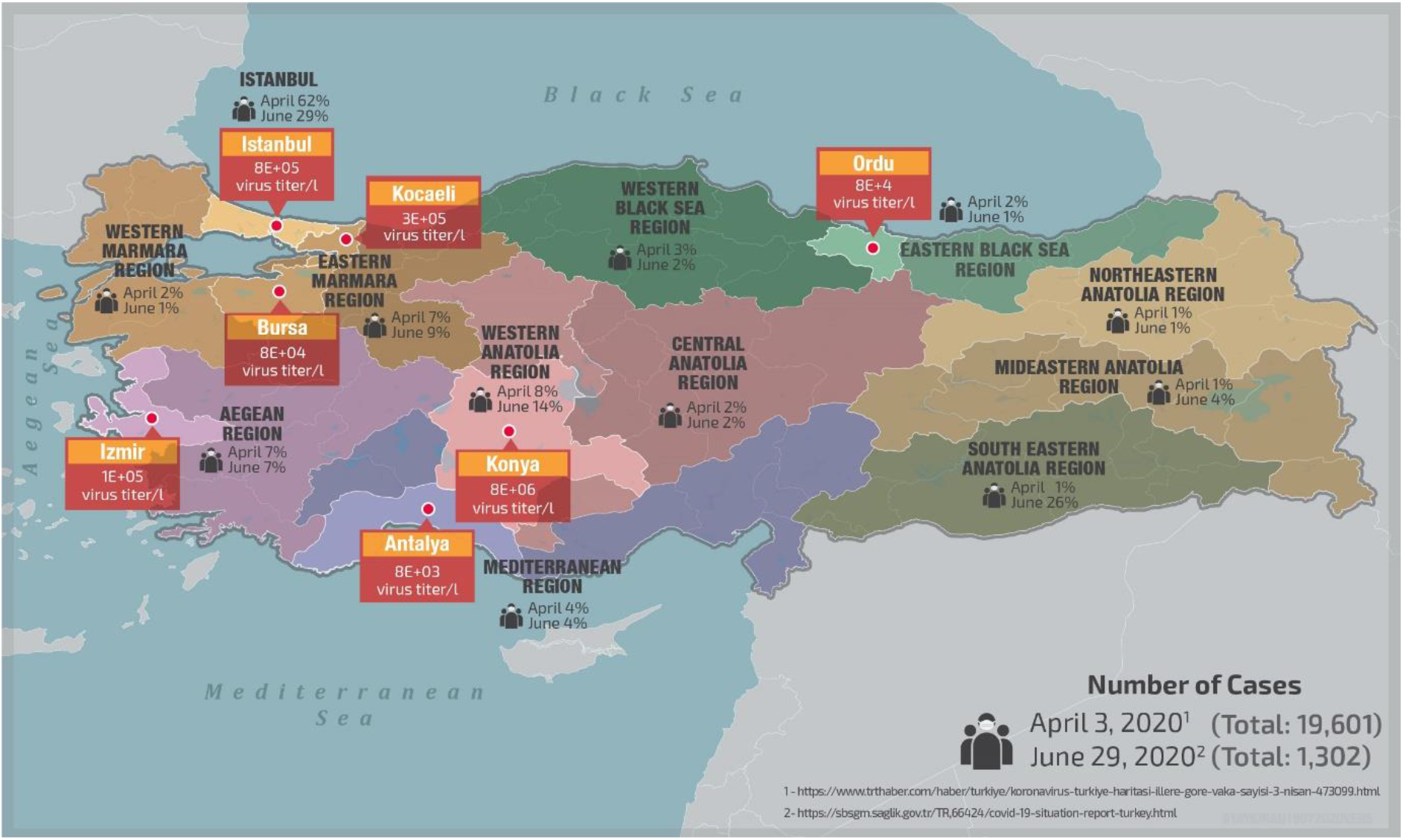
Overall results of SARS-CoV-2 surveillance in wastewater and sludges of Turkey from May 2020 to June 2020

In mega city Istanbul (Fig. 13), SARS-CoV-2 was detected more concentrated in the effluent samples with respect to the influent samples. Moreover, The SARS-CoV-2 detection in the waste activated sludge samples was about 14 % more with respect to the influent samples. These observations might be due to adsorption of SARS-CoV-2 on sludge and release of adsorbed viruses with time as the waste activated sludge was recycled for days in the treatment plant due to long sludge age in extended aeration systems, common in Istanbul WWTPs.

**Fig 13.**
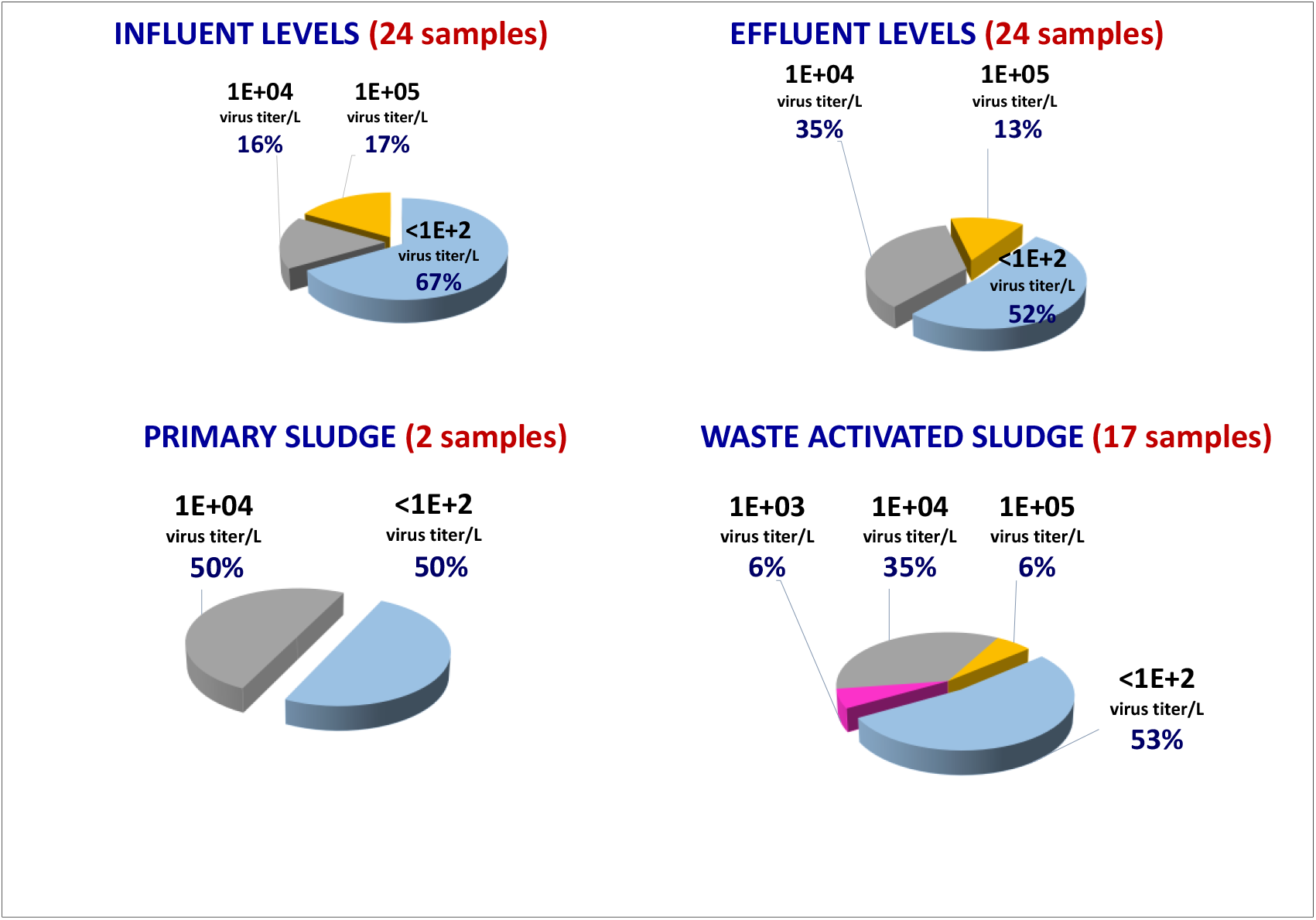
Distribution of SARS-CoV-2 titers in Istanbul influent, effluent and sludge samples from May 2020 to June 2020

All throughout the WWTPs in Turkey (Fig.14), no significant differences were observed among influent, effluent and sludge measurements. Titers of SARS-CoV-2 have been detected ranging from 2.7E+3 to 8.2E+6 for the influent samples, 6.0E+3 to 7.6E+6 for the effluent samples, 1.6E+4 to1.1E+6 for the primary sludge samples and 2.6E+2 to 2.8E+6 for the waste activated sludge samples.

**Fig 14.**
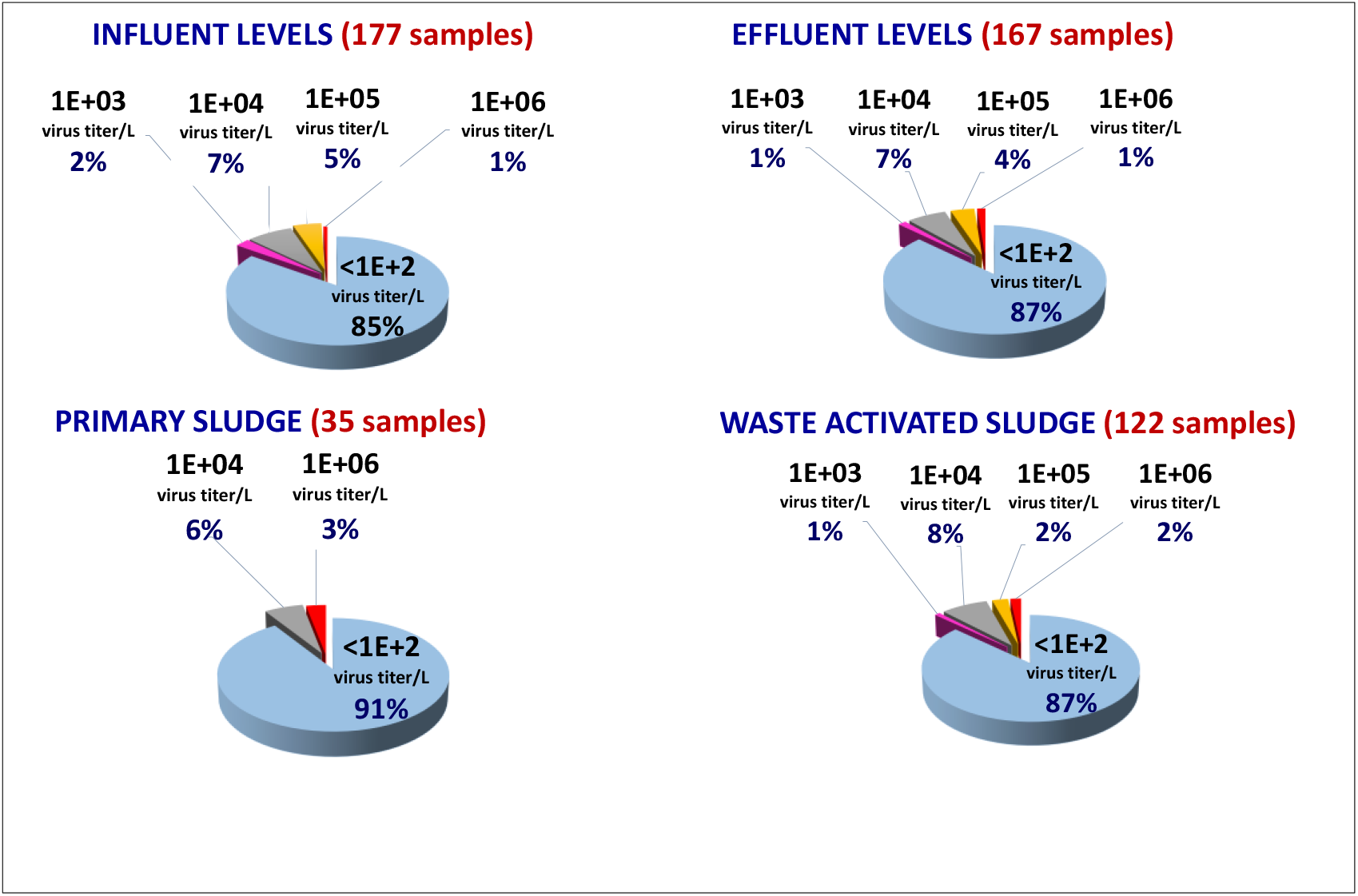
Distribution of SARS-CoV-2 titers in influent, effluent and sludge samples of 81 cities in Turkey from May 2020 to June 2020

### 4.2. SARS-CoV-2 Activity Test Results

Influent, effluent, primary sludge and waste activated sludge samples of Istanbul, Izmir, Izmit, Bursa and Konya cities demonstrating positive results of SARS-CoV-2 were tested in order to evaluate the possible activity of virus in treatment plants. Among the 31 positive samples tested, only in 1 waste activated sludge sample of Konya WWTP the SARS-CoV-2 virus was found active.

### 4.3. Comments on Negative qPCR results

As mentioned in Section 4.1, among the 81 cities scanned from May 2020 to June 2020, SARS-CoV-2 was only detected in 7 cities wastewater and sludge samples. All these tests were performed using qPCR primers targeting RdRp gene (Section 3.3) since initial trials with the primer targeting N1N2 were not successful to detect SARS-CoV-2. However, starting from June 2020, SARS-CoV-2 could not be detected in any city, although there were cities like Istanbul reporting significant number of cases at that time. Hence, in September 2020, it was decided to change primer targeting RdRp gene with the primer targeting N1N2 gene. Using the primer targeting N1N2 gene, we again started to monitor SARS-CoV-2 in the samples. The parallel readings conducted with primers targeting RdRp gene were still negative.

## Data Availability

ll data referred to in the manusrpt are available

## Ethics Statement

The work did not involve any human subject and animal experiments.

## Acknowledgments

This work was financed by Republic of Turkey, Ministry of Agriculture and Forestry.

The authors wish to acknowledge the Turkish Water Institute (SUEN) for the coordination and execution of this study. We wish to express our appreciation to the Ministry’s Istanbul Pendik and Samsun Veterinary Control Central Research Institute for their hard work and rapid analysis of the wastewater samples. We thank to DSI (State Hydraulic Works) for their logistic support for intercity sample transportation. We also thank to all municipalities for their cooperation and efforts to collect and preserve the sewage samples rapidly. Our special thanks to Env. Eng. Salim Yaykiran from SUEN for regional maps and Env.Eng. Sumeyye Celik from Marmara University her contribution to gather and collate up to date information about the worldwide studies on SARS-CoV-2 in wastewater.

